# COVID-19 Lockdown in a Kenyan Informal Settlement: Impacts on Household Energy and Food Security

**DOI:** 10.1101/2020.05.27.20115113

**Authors:** Matthew Shupler, James Mwitari, Arthur Gohole, Rachel Anderson de Cuevas, Elisa Puzzolo, Iva Čukić, Emily Nix, Dan Pope

**Affiliations:** Department of Public Health and Policy, University of Liverpool, Liverpool, United Kingdom; School of Public Health, Amref International University, Nairobi, Kenya; Global LPG Partnership (GLPGP), 654 Madison Avenue, New York, United States

**Keywords:** Clean cooking, household energy, COVID-19, air pollution, Nairobi, informal urban settlement, food security

## Abstract

A COVID-19 lockdown may impact household fuel use and food security for ∼700 million sub-Saharan Africans who rely on polluting fuels (e.g. wood, kerosene) for household energy and typically work in the informal economy. In an informal settlement in Nairobi, surveys administered before (n=474) and after (n=194) a mandatory COVID-19-related community lockdown documented socioeconomic/household energy impacts. During lockdown, 95% of participants indicated income decline or cessation and 88% reported being food insecure. Three quarters of participants cooked less frequently and half altered their diet. One quarter (27%) of households primarily using liquefied petroleum gas (LPG) for cooking before lockdown switched to kerosene (14%) or wood (13%). These results indicate the livelihoods of urban Kenyan families were deleteriously affected by COVID-19 lockdown, with a likely rise in household air pollution from community-level increases in polluting fuel use. To safeguard public health, policies should prioritize enhancing clean fuel and food access among the urban poor.

## INTRODUCTION

Approximately 2.8 billion people worldwide use polluting fuels, including solid fuels (e.g. wood, charcoal) and kerosene, for household cooking, heating and lighting.^1^ In sub-Saharan Africa (SSA), HAP was attributed to 9% of mortality and 7% of underlying morbidity in 2018.^2^ Amid the coronavirus disease 2019 (COVID-19) pandemic, there is concern about increased disease severity in SSA due to higher baseline prevalence of infectious diseases (e.g. tuberculosis, pneumonia)^3^ and non-communicable respiratory/cardiovascular diseases,^4–7^ which are all causally linked to HAP exposure.^8–10^

To combat the adverse health, environmental and social effects of reliance on polluting fuels and associated HAP exposures,^11,12^ several African countries (e.g. Kenya, Ghana, Cameroon), have set aspirational targets for rapid market expansion of liquefied petroleum gas (LPG) for cooking.^13,14^ Despite being a fossil fuel, LPG is considered ‘clean’ because it burns very efficiently, emitting no black carbon and low levels of fine particulate matter (PM_2.5_)^15,16^ and contributes to forest protection.^17,18^ LPG is a clean energy alternative actively promoted in SSA, with several consumer-tailored solutions, including consumer-finance mechanisms (e.g. pay-asyou-go (PAYG), mobile payments) being evaluated to increase its affordability.^19^ However, national scale-up efforts may become increasingly difficult because of the financial repercussions of the COVID-19 pandemic, government-mandated community lockdowns to mitigate its spread and logistical/trade constraints affecting supply/distribution.^20^ With affordability of LPG being a critical barrier to uptake in SSA before the pandemic,^21,22^ income loss during lockdown can further diminish its scalability, and increase levels of food insecurity.^23^ A World Food Programme report predicts a doubling of hunger risk from the COVID-19 pandemic.^24^

In Africa, up to 90% of urban development consists of informal settlements.^25^ These settlements suffer from overcrowding, with limited water/sanitation facilities and cooking typically done in a single, multipurpose room. Household energy sources can be variable in these environments outside the context of a pandemic; cooking fuels may be routinely ‘stacked’ (combination of clean and/or polluting fuels)^26,27^ or primary fuels switched from clean to polluting fuels (‘reverse switching’) due to unexpected/seasonal changes in income.^28,29^ Thus, financial hardship brought about by a mandatory lockdown can potentially spur community-level reverse switching, leading to household PM_2.5_ level increases above the World Health Organization (WHO) Indoor Air Quality guidelines,^30,31^ which can be intensified by tightly-packed housing, as smoke can infiltrate neighboring homes.^32^ A lockdown can therefore have severe negative socioeconomic and health consequences,^33,34^ with limited success in mitigating COVID-19 spread in these densely populated areas,^35^ where cramped conditions and shared facilities make physical distancing infeasible.

In Kenya, a mandatory lockdown was instated on March 25, 2020, two weeks after the first recorded case of COVID-19, followed by a dusk-to-dawn curfew (7PM-5AM) instituted on April 7, 2020. With 56% of the urban Kenyan population living in informal settlements (>2 million living in Nairobi),^36^ the effects of an unprecedented community lockdown on household energy decisions, food security and air pollution exposures of the urban poor are uncertain. This pre-post study provides valuable information on impacts of the COVID-19 lockdown among a vulnerable population living in Mukuru Kwa Reuben informal settlement in Nairobi.

## METHODS

The study setting of Mukuru Kwa Rueben is an informal settlement with over 500,000 residents along the Nairobi Ngong river, situated on polluted lands in the industrial area of Nairobi. A population-based survey on socioeconomic factors and fuel use patterns was administered to the main cook (or another member of the household, if unavailable) via door-to-door sampling from December 2019-March 2020 (before the lockdown went into effect). Field workers that administered the surveys also serve as community health volunteers in kwa Rueben and had an earned level of trust among study participants. Questions about familiarity with pay-as-you-go LPG (PAYG) consumer finance mechanisms were asked to a subset of participants; a sensitivity analysis was conducted to compare socioeconomic characteristics between the PAYG sample and full sample via Pearson’s chi-squared tests (categorical data) and two-sample t-tests (continuous data) for independence. All survey data was collected via smartphones and securely transferred to an online storage system using Mobenzi Research, a secure digital platform that has been used successfully in previous health monitoring studies.^48,49^

After Nairobi went into lockdown on March 20, 2020, a telephone-based survey was conducted from April 20–30 among consenting participants from the baseline survey, to document post-lockdown impacts on food security and cooking behaviours. A sensitivity analysis was conducted to compare demographics among the baseline and follow-up sample in the pre-post study using the same statistical tests as above.

Additionally, real-time ambient PM_2.5_ levels before and during the lockdown period were obtained from an open-source sensor network (https://sensors.africa/) that consists of over 70 low-cost, laser scattering monitors (Nova particulate matter sensor SDS011) deployed throughout cities in Kenya, Tanzania and Nigeria. The project is seed-funded by *innovateAFRICA* (not-for-profit consultancy). Daily average PM_2.5_ concentrations were examined from sensors in Nairobi located near Mukuru kwa Rueben (four sensors operational in March/April 2020 and three operational in April 2019).

Ethical approval for this research was obtained from the University of Liverpool in Liverpool, United Kingdom and Amref Health Africa in Nairobi, Kenya. Generation of figures and all analyses were conducted in R version 3.5.1.^50^

## RESULTS

### Baseline Demographics

A total of 474 randomly selected participants living in Mukuru Kwa Reuben completed a baseline survey (before lockdown), of whom 88% (n=419) were the main cook of the household. The mean age was 30 years old, and 70% of respondents were female. Two-thirds of respondents had a monthly household income less than or equal to 15,000 Kenyan Shilling (Ksh) (∼$140 USD). Almost half (43%) of households experienced seasonal fluctuations in their income. Nine out of ten households comprised one or two rooms (Table S1). Three-quarters (77%) of respondents reported having access to drinking water in their home and nearly all (97%) used a communal standpipe as their main water source.

### Household Energy Patterns Before Lockdown

Among baseline survey respondents, half (49%; n=232) used LPG as their primary fuel, 44% (n=207) used kerosene and the few remaining households used charcoal/charcoal briquettes (4%; n=15), electricity (2%; n=7) or wood (1%; n=4) (Figure 1-left). A quarter (26%) of households stacked two or more fuels, with the most common fuel combination being LPG (primary) and kerosene (secondary) (8% of households; n=38) (Figure 1-right).

**Figure 1.**
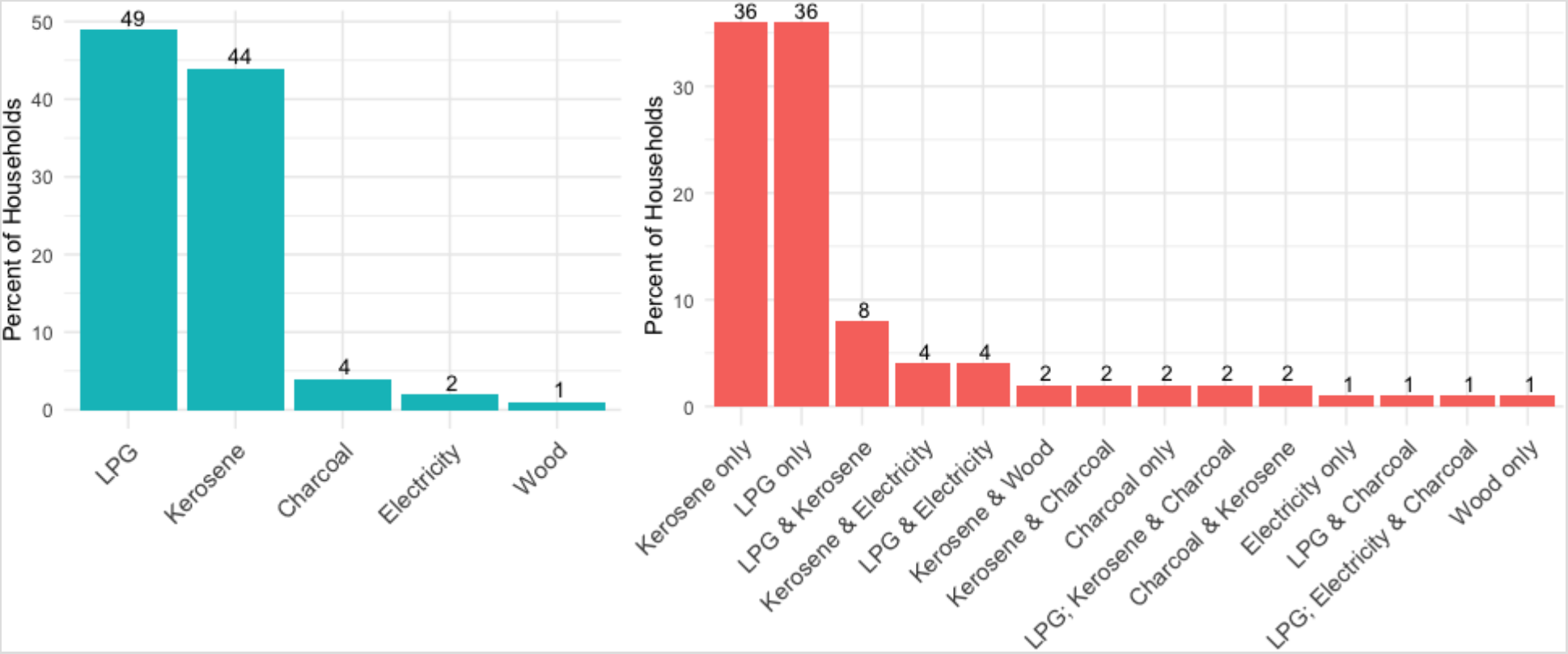
(left). Baseline prevalence of primary fuels in Mukuru Kwa Reuben informal urban settlement. (right) Baseline prevalence of primary, secondary and tertiary cooking fuel combinations. Fuels listed in order of primary, secondary and tertiary usage.

Four out of five of households (83%) cooking with LPG at baseline started using it less than two years earlier (see Supplemental Information; Table S9). Over half (59%; N=127) of households not cooking with LPG had used it in the past, and > 90% of these households expressed an interest in cooking with LPG (Table S10).

Nearly all (90%) families cooked inside the home in a single room. Households cooking with LPG self-reported a weekday mean cooking time (2.9 hours) 1.5 hours less than that of wood-using households (4.5 hours) (Table 1). Only 2% of respondents obtained their primary cooking fuel for free. The frequency of fuel purchases among households paying for their cooking fuel varied dramatically by primary fuel type; almost all LPG primary users (95%) buying refills (85% of households used 6 kilogram cylinders (Table S9)) once a month or every 2–4 months. Conversely, two-thirds of participants primarily cooking with kerosene (66%) and over half using charcoal (57%) purchased their fuels daily (Table 1). Despite self-reported monthly mean LPG fuel costs being the least expensive ($1267 Ksh/month (∼$12 USD/month or $0.40 USD/day)) when compared to all polluting fuel types, the mean cost of LPG per single purchase was highest because it was acquired in monthly increments (Table 1). Almost half (45%) of participants using kerosene as a primary fuel perceived the cost of LPG to be expensive or very expensive, compared with 21% of LPG users. One quarter (25%) of kerosene primary fuel users perceived LPG to be dangerous or very dangerous, compared with only 3% of LPG users (Table 1).

**Table 1.**
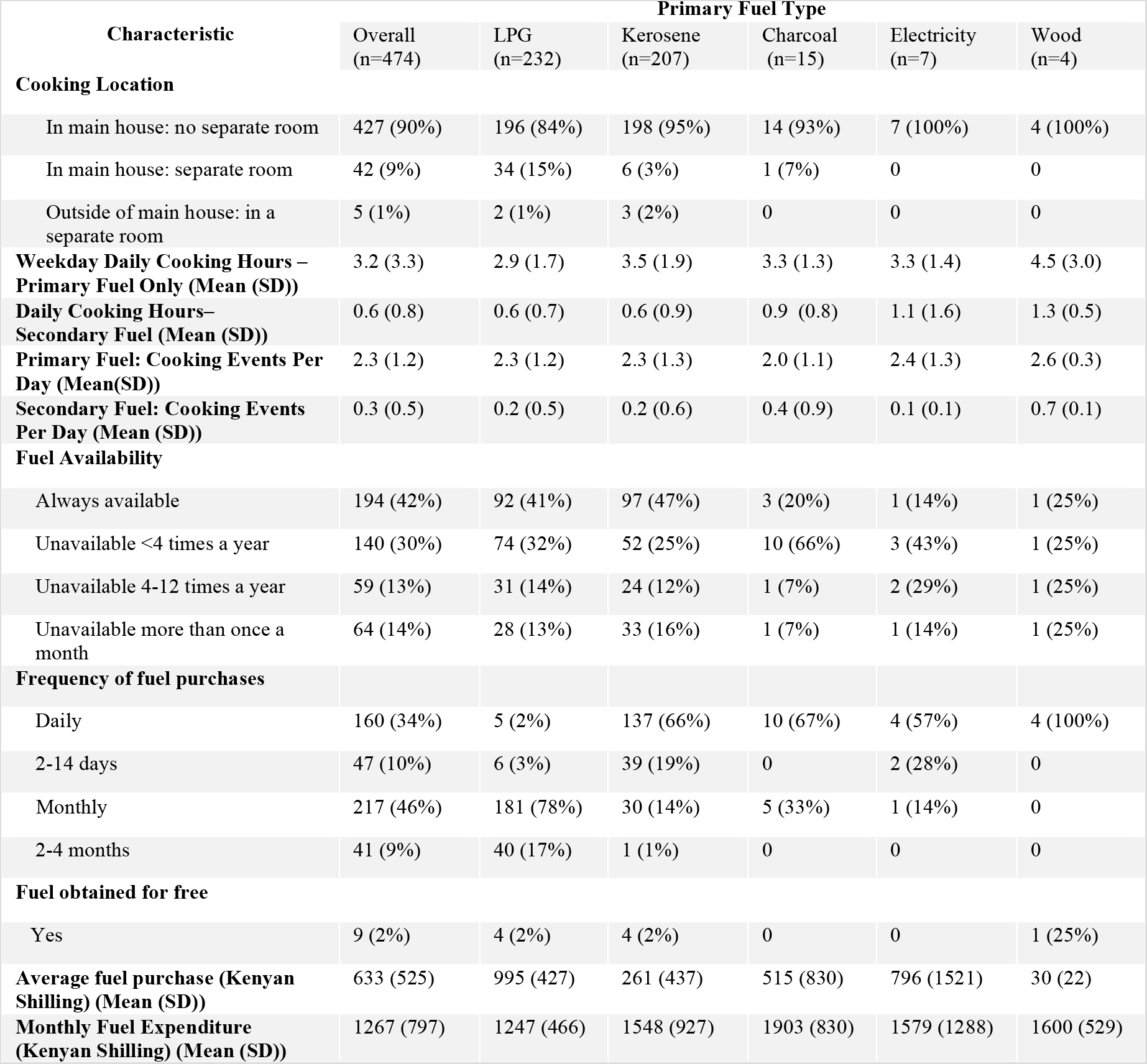
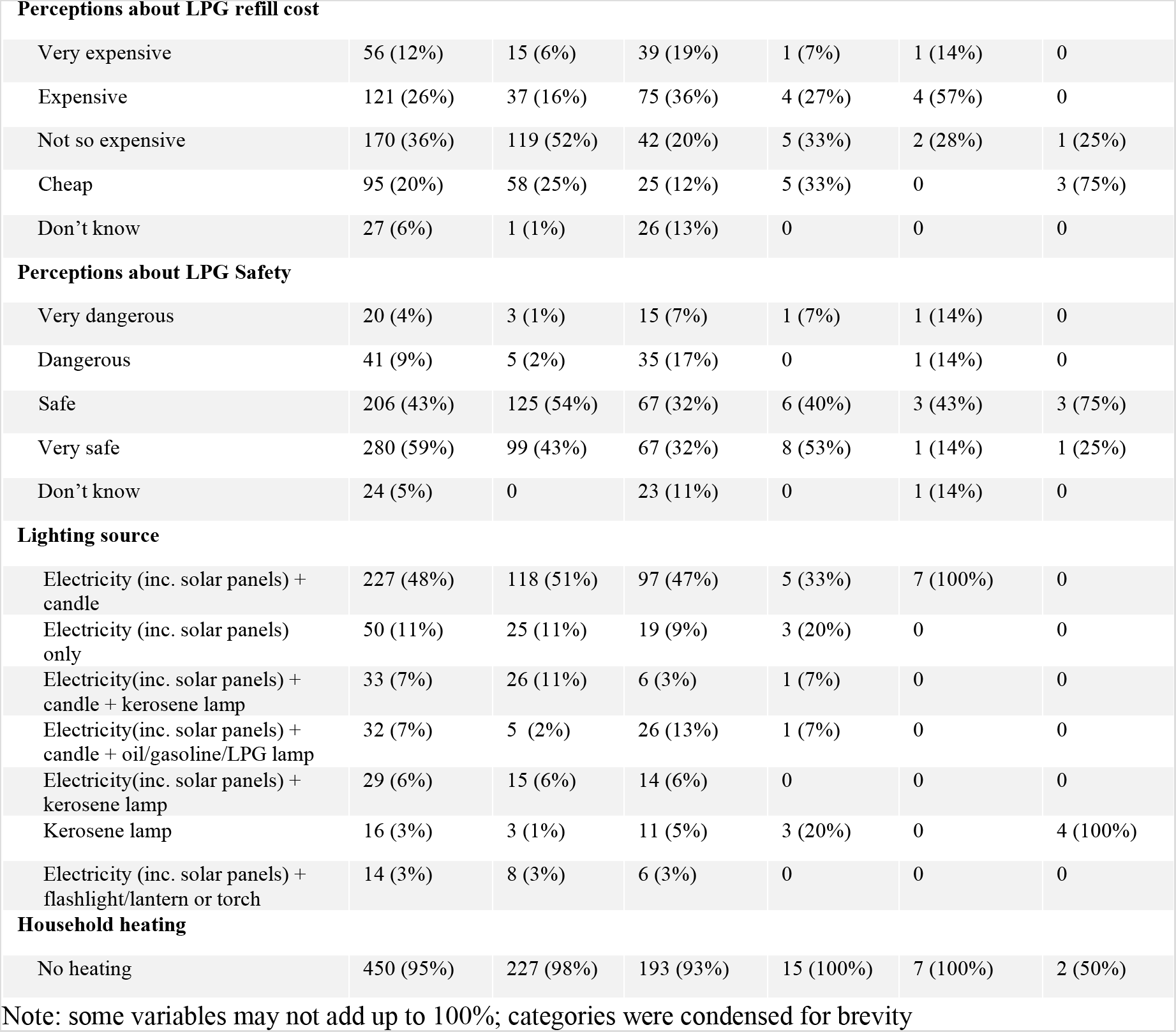
Cooking characteristics by primary fuel type.

| Characteristic | Primary Fuel Type |  |  |  |  |  |
| --- | --- | --- | --- | --- | --- | --- |
|  | Overall<br>(n=474) | LPG<br>(n=232) | Kerosene<br>(n=207) | Charcoal<br>(n=15) | Electricity<br>(n=7) | Wood<br>(n=4) |
| **Cooking Location** |  |  |  |  |  |  |
| In main house: no separate room | 427 (90%) | 196 (84%) | 198 (95%) | 14 (93%) | 7 (100%) | 4 (100%) |
| In main house: separate room | 42 (9%) | 34 (15%) | 6 (3%) | 1 (7%) | 0 | 0 |
| Outside of main house: in a separate room | 5 (1%) | 2 (1%) | 3 (2%) | 0 | 0 | 0 |
| **Weekday Daily Cooking Hours – Primary Fuel Only (Mean (SD))** | 3.2 (3.3) | 2.9 (1.7) | 3.5 (1.9) | 3.3 (1.3) | 3.3 (1.4) | 4.5 (3.0) |
| **Daily Cooking Hours– Secondary Fuel (Mean (SD))** | 0.6 (0.8) | 0.6 (0.7) | 0.6 (0.9) | 0.9 (0.8) | 1.1 (1.6) | 1.3 (0.5) |
| **Primary Fuel: Cooking Events Per Day (Mean(SD))** | 2.3 (1.2) | 2.3 (1.2) | 2.3 (1.3) | 2.0 (1.1) | 2.4 (1.3) | 2.6 (0.3) |
| **Secondary Fuel: Cooking Events Per Day (Mean (SD))** | 0.3 (0.5) | 0.2 (0.5) | 0.2 (0.6) | 0.4 (0.9) | 0.1 (0.1) | 0.7 (0.1) |
| **Fuel Availability** |  |  |  |  |  |  |
| Always available | 194 (42%) | 92 (41%) | 97 (47%) | 3 (20%) | 1 (14%) | 1 (25%) |
| Unavailable <4 times a year | 140 (30%) | 74 (32%) | 52 (25%) | 10 (66%) | 3 (43%) | 1 (25%) |
| Unavailable 4-12 times a year | 59 (13%) | 31 (14%) | 24 (12%) | 1 (7%) | 2 (29%) | 1 (25%) |
| Unavailable more than once a month | 64 (14%) | 28 (13%) | 33 (16%) | 1 (7%) | 1 (14%) | 1 (25%) |
| **Frequency of fuel purchases** |  |  |  |  |  |  |
| Daily | 160 (34%) | 5 (2%) | 137 (66%) | 10 (67%) | 4 (57%) | 4 (100%) |
| 2-14 days | 47 (10%) | 6 (3%) | 39 (19%) | 0 | 2 (28%) | 0 |
| Monthly | 217 (46%) | 181 (78%) | 30 (14%) | 5 (33%) | 1 (14%) | 0 |
| 2-4 months | 41 (9%) | 40 (17%) | 1 (1%) | 0 | 0 | 0 |
| **Fuel obtained for free** |  |  |  |  |  |  |
| Yes | 9 (2%) | 4 (2%) | 4 (2%) | 0 | 0 | 1 (25%) |
| **Average fuel purchase (Kenyan Shilling) (Mean (SD))** | 633 (525) | 995 (427) | 261 (437) | 515 (830) | 796 (1521) | 30 (22) |
| **Monthly Fuel Expenditure (Kenyan Shilling) (Mean (SD))** | 1267 (797) | 1247 (466) | 1548 (927) | 1903 (830) | 1579 (1288) | 1600 (529) |
| **Perceptions about LPG refill cost** |  |  |  |  |  |  |
| Very expensive | 56 (12%) | 15 (6%) | 39 (19%) | 1 (7%) | 1 (14%) | 0 |
| Expensive | 121 (26%) | 37 (16%) | 75 (36%) | 4 (27%) | 4 (57%) | 0 |
| Not so expensive | 170 (36%) | 119 (52%) | 42 (20%) | 5 (33%) | 2 (28%) | 1 (25%) |
| Cheap | 95 (20%) | 58 (25%) | 25 (12%) | 5 (33%) | 0 | 3 (75%) |
| Don't know | 27 (6%) | 1 (1%) | 26 (13%) | 0 | 0 | 0 |
| **Perceptions about LPG Safety** |  |  |  |  |  |  |
| Very dangerous | 20 (4%) | 3 (1%) | 15 (7%) | 1 (7%) | 1 (14%) | 0 |
| Dangerous | 41 (9%) | 5 (2%) | 35 (17%) | 0 | 1 (14%) | 0 |
| Safe | 206 (43%) | 125 (54%) | 67 (32%) | 6 (40%) | 3 (43%) | 3 (75%) |
| Very safe | 280 (59%) | 99 (43%) | 67 (32%) | 8 (53%) | 1 (14%) | 1 (25%) |
| Don't know | 24 (5%) | 0 | 23 (11%) | 0 | 1 (14%) | 0 |
| **Lighting source** |  |  |  |  |  |  |
| Electricity (inc. solar panels) + candle | 227 (48%) | 118 (51%) | 97 (47%) | 5 (33%) | 7 (100%) | 0 |
| Electricity (inc. solar panels) only | 50 (11%) | 25 (11%) | 19 (9%) | 3 (20%) | 0 | 0 |
| Electricity(inc. solar panels) + candle + kerosene lamp | 33 (7%) | 26 (11%) | 6 (3%) | 1 (7%) | 0 | 0 |
| Electricity(inc. solar panels) + candle + oil/gasoline/LPG lamp | 32 (7%) | 5 (2%) | 26 (13%) | 1 (7%) | 0 | 0 |
| Electricity(inc. solar panels) + kerosene lamp | 29 (6%) | 15 (6%) | 14 (6%) | 0 | 0 | 0 |
| Kerosene lamp | 16 (3%) | 3 (1%) | 11 (5%) | 3 (20%) | 0 | 4 (100%) |
| Electricity (inc. solar panels) + flashlight/lantern or torch | 14 (3%) | 8 (3%) | 6 (3%) | 0 | 0 | 0 |
| **Household heating** |  |  |  |  |  |  |
| No heating | 450 (95%) | 227 (98%) | 193 (93%) | 15 (100%) | 7 (100%) | 2 (50%) |

Note: some variables may not add up to 100%; categories were condensed for brevity

Approximately 95% of households used electricity for lighting, but only 11% used it exclusively (Table 2). Households without electricity mostly used kerosene lamps as the main lighting source (3%). Nearly half of electrified households used candles in times of power cuts, while other households also resorted to kerosene lamps (13%), oil/gasoline/LPG lamps (7%) or flashlights/lanterns/torches (3%). Only 5% of sampled households ever heat their homes, with most (81%) only for 4 months or less annually.

**Table 2.**
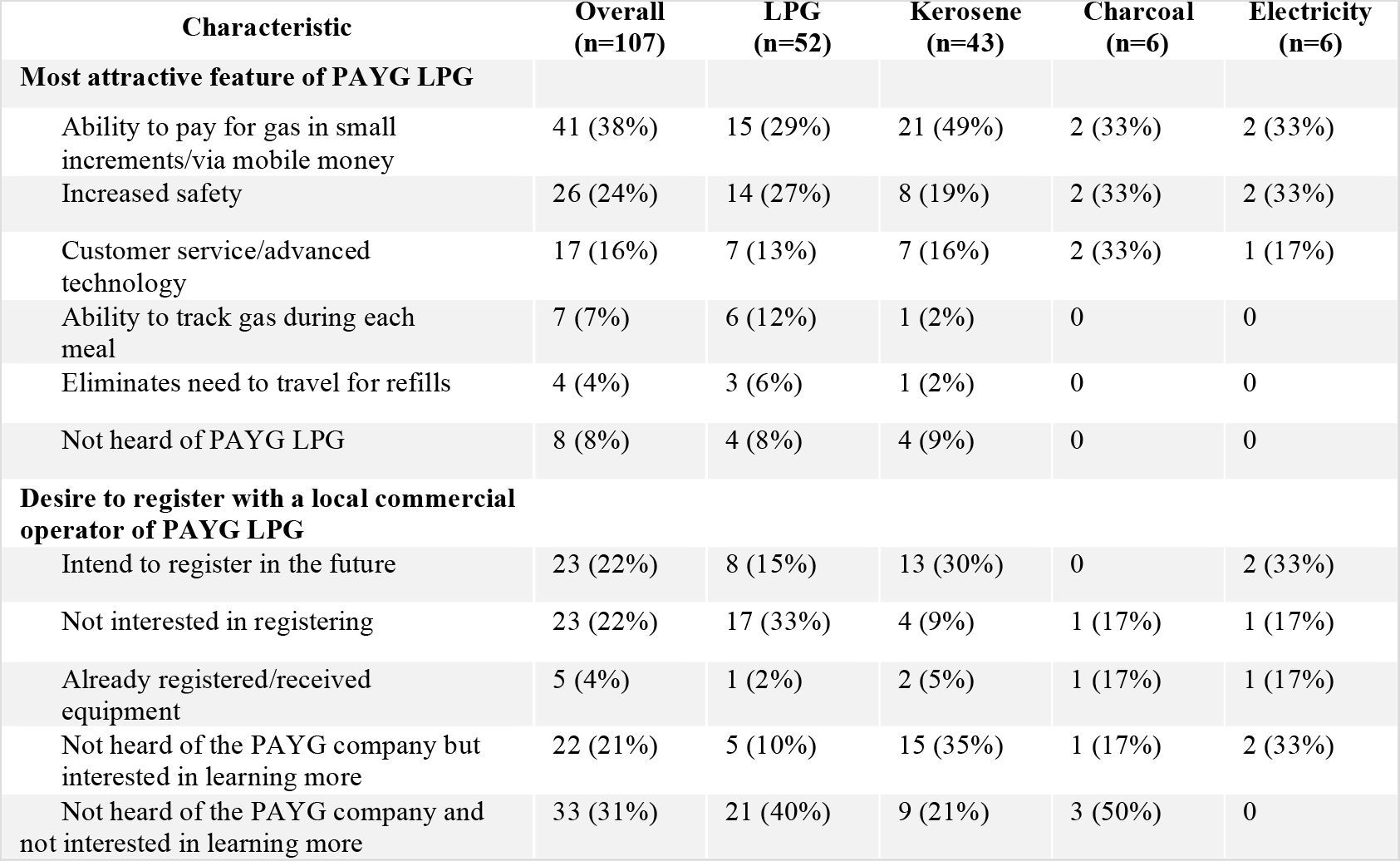
Interest in Pay-As-You-Go (PAYG) LPG smart-meter technology (n = 107)

#### Pay-As-You-Go Liquefied Petroleum Gas (Before Lockdown)

Due to increasing interest in Kenya about the potential for pay-as-you-go (PAYG) LPG smart-meter technology^19^ to transition individuals to LPG for cooking, a subset of 107 participants (similar in demographic profile to the full baseline sample (Table S3)) were questioned before the lockdown about their perceptions of PAYG LPG technology. The most attractive reported feature was ability to pay for gas in small amounts (37%), followed by increased safety (27%) and customer service (14%) (Table 2). Elimination of the need to travel for gas refills was not a highly motivating feature (4%).

Half (52%) of respondents had not heard of PAYG LPG; the remaining half were equally split between those intending and not intending to register (22% each) for the equipment, and 4% of respondents had already registered or were existing customers (Table 2). Interest in registering for PAYG technology among households already using LPG as a primary fuel (15%) was half that of households currently using kerosene (30%). The average monthly household income among the 23 participants not interested in registering for PAYG LPG was higher than the average of the overall study population, with approximately 40% earning a monthly income greater than 15,000 Ksh ($140 USD), compared to 22% among the full sample (Table S1).

#### Changes in Household Characteristics During Lockdown

The sampling frame for follow up telephone-based surveys administered during the COVID-19 lockdown in April 2020, consisted of 60% (n=285) of baseline respondents consenting to be contacted again. The final analytic sample consisted of 194/285 participants (41% of original baseline survey respondents). A lower-than-expected rate of follow up was primarily due to participants’ mobile being switched off (likely because of inability to pay mobile phone bills amidst a time of financial hardship during the pandemic, and lack of power connectivity for charging among those who had travelled to rural villages). No difference was observed in demographic profile between the 474 baseline and 194 follow-up participants (Table S2). Nearly all participants (95%) reported decreases in income during lockdown, with (88%) indicating that their decreased income was insufficient to purchase food needed by the household (Table 3); a third of households (34%) indicated total cessation of income amid the lockdown. Over half of households (56%) cooked less frequently while in lockdown, while 6% cooked more frequently. The change in cooking frequency varied by primary fuel type with 24% of households using wood cooking more frequently compared with only 6% of households using LPG and 3% using kerosene (Table S4).

**Table 3.**
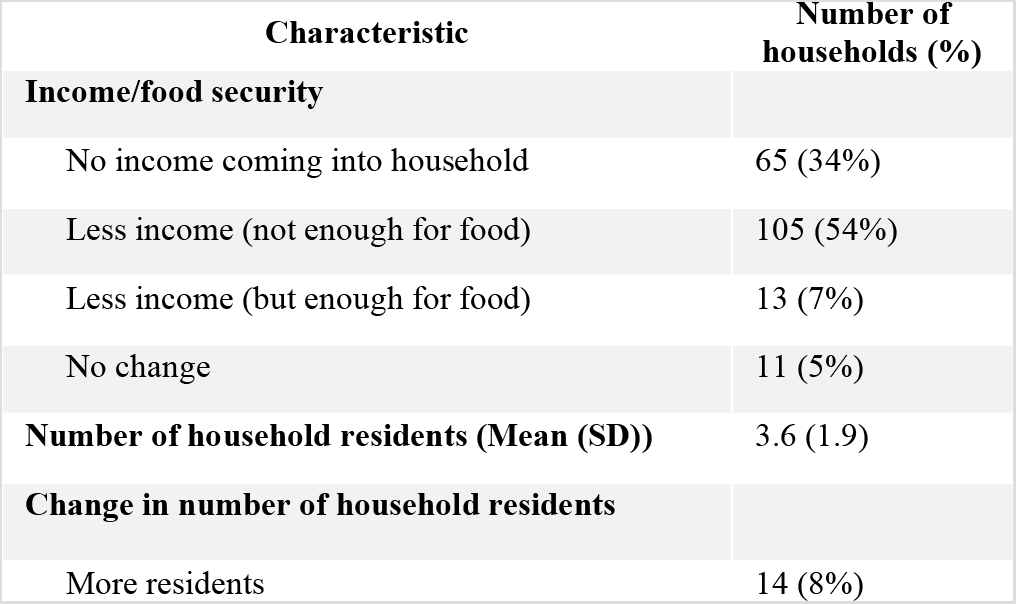
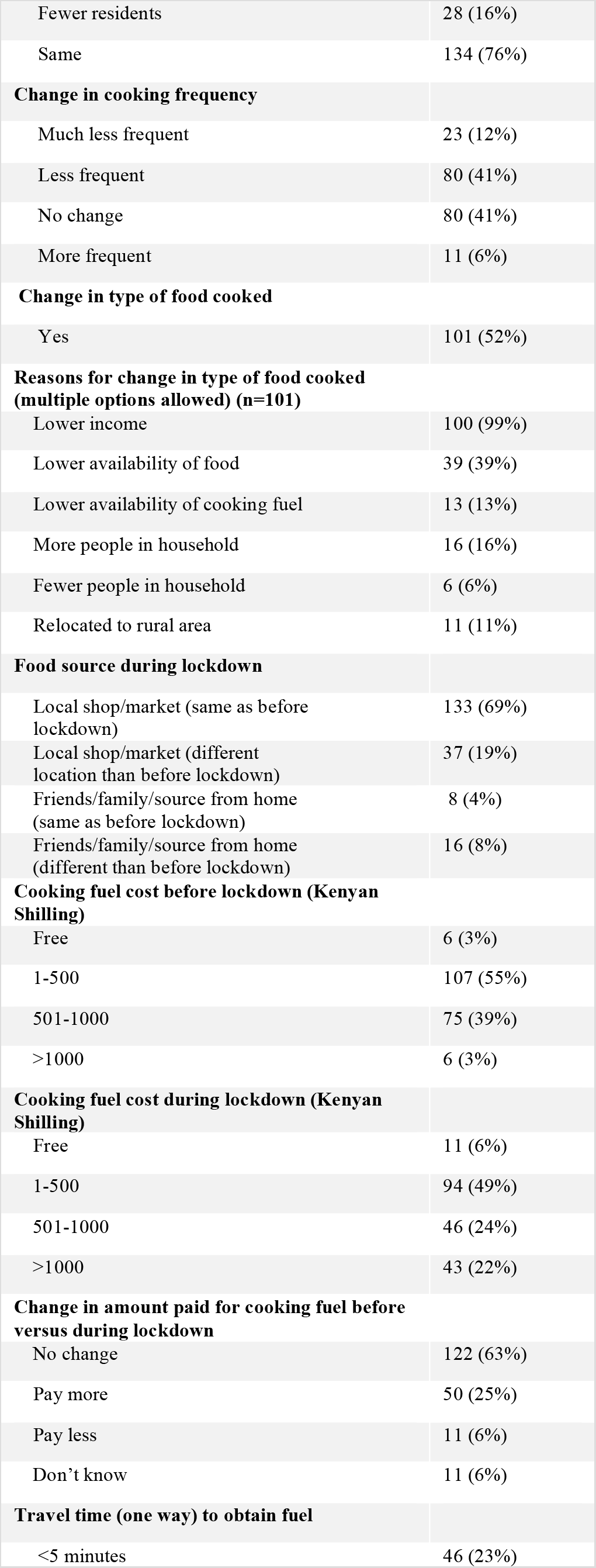
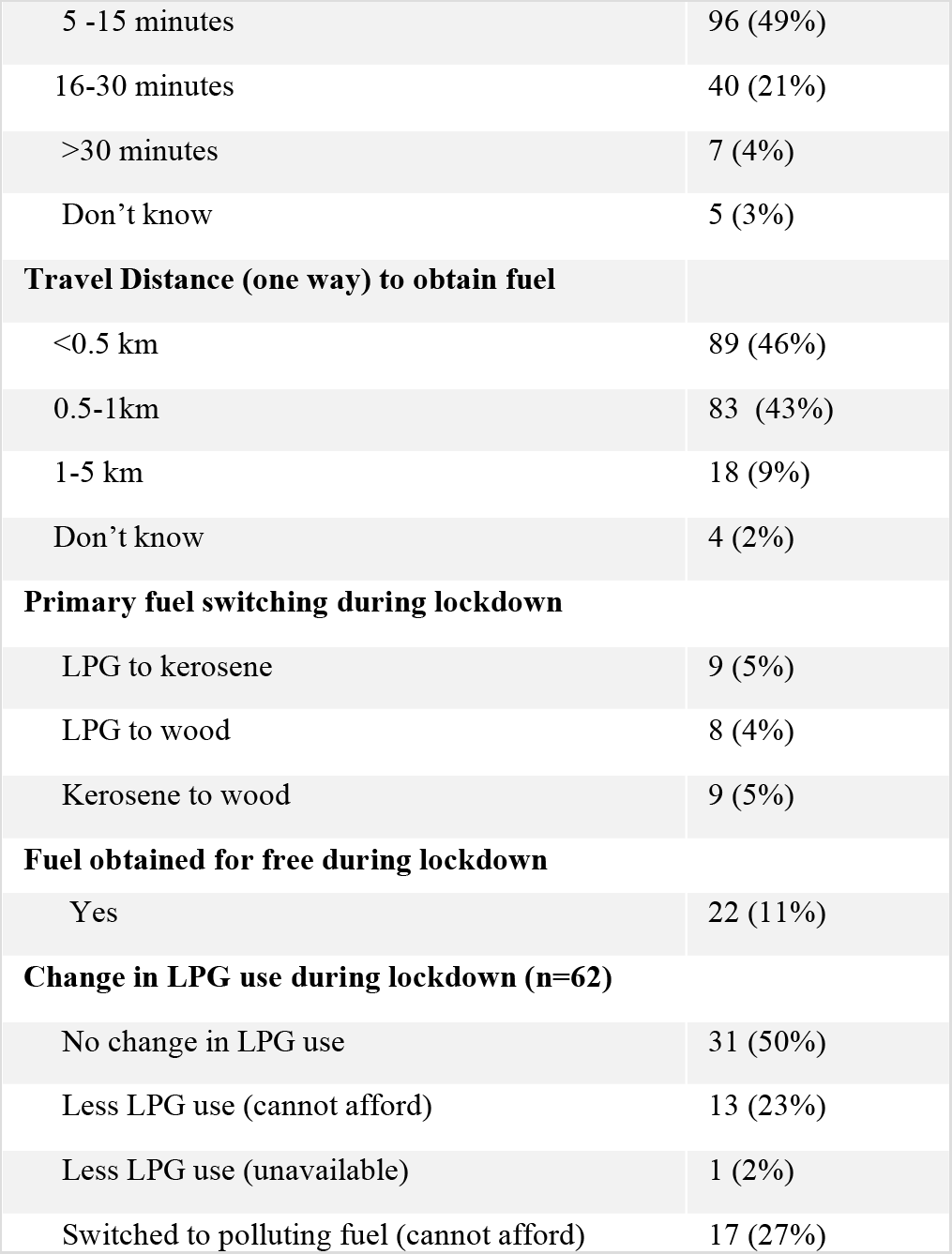
Effect of COVID-19 lockdown on livelihoods in Mukuru Kwa Rueben informal urban settlement (n = 194)

Half (52%) of participants indicated a change in the type of food they cook. Common dietary changes were reductions in meat/fish, milk/milk tea and bread/chapati, with higher consumption of vegetables (Table S4). In addition to insufficient income, lower food availability (39%) and increases (16%) or decreases (8%) in the number of household inhabitants during lockdown were main reasons alterations in dietary behaviour. A quarter (27%) of households switched their main food vendor/retailer during the lockdown, with 8% of households resorting to farming/livestock as their new, main food source.

One quarter of participants (25%) paid more for their primary cooking fuel during lockdown than before the pandemic (Table 3). The percentage of households using LPG that paid a higher rate after lockdown (55%) was three times higher than the proportion of households using kerosene that paid a higher rate after the lockdown mandate (18%) (Table S7). More specifically, the percent of households using LPG paying a total amount of > 1,000 Ksh for refills increased dramatically (55%) from 4% at baseline to 59% during lockdown, compared with only an 8% rise (3% to 11%) in the proportion among kerosene users (Table S8). Despite fuel cost increases, availability was not an issue experienced by participants during lockdown; 73% of participants obtained their fuel in under 15 minutes (one way) travel time.

Primary fuel switching occurred among 14% (n=27) of households in response to the lockdown, with LPG users (26%) being three times more likely to switch than kerosene users (8%). Previous LPG users switched to kerosene (n=9) or wood (n=8), and households previously using kerosene switched to wood (n=9) (Figure 2-left). The prevalence of wood as a primary fuel increased by a factor of five (2% to 11%) during lockdown, in conjunction with a 9% decline in primary LPG use for cooking (34% to 25%) (Figure 2-right). Further, approximately a third (31%) of participants that continued to use LPG during the lockdown reported less use (Table 3).

**Figure 2.**
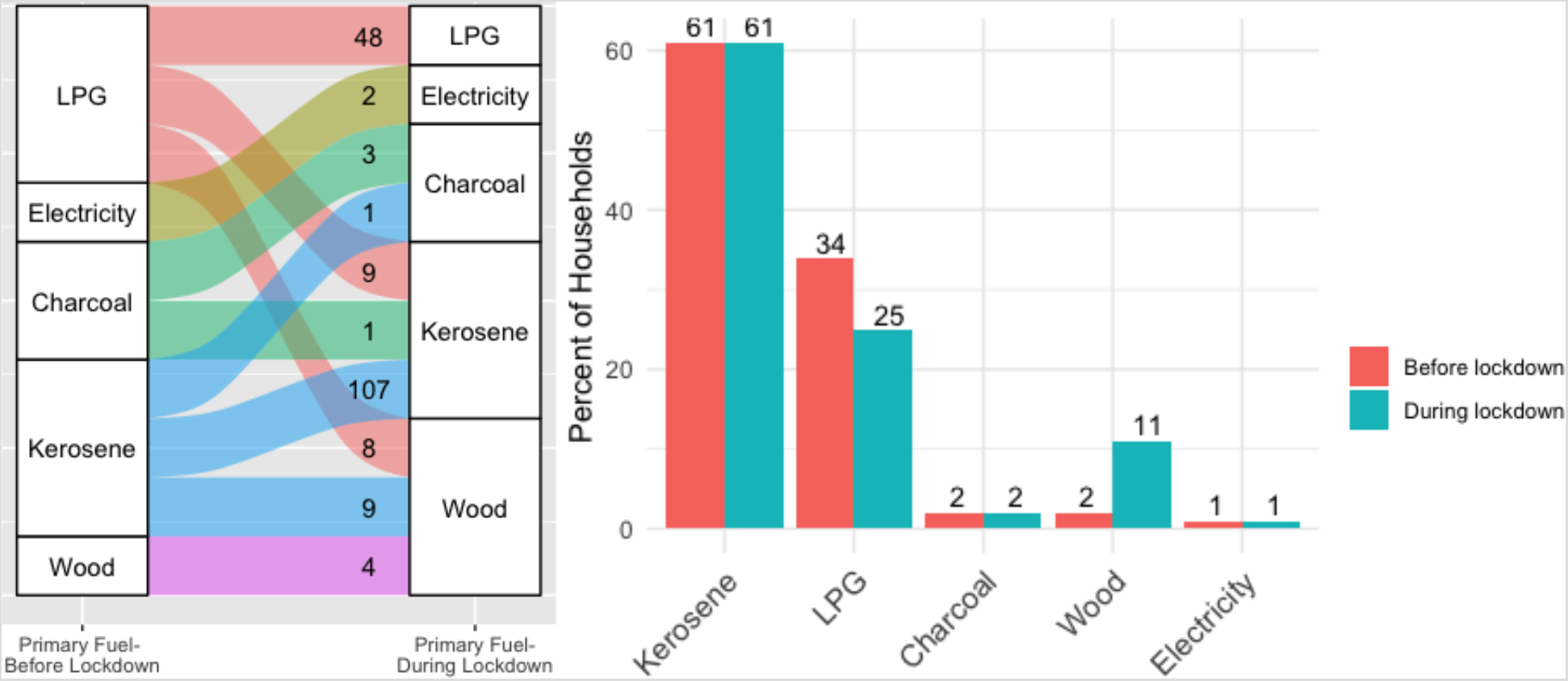
(left). Number of households switching between different primary fuel types during lockdown. (right) Prevalence (%) of primary fuel types in Mukuru Kwa Rueben informal urban settlement before and during lockdown (n=194)

#### Ambient Air Pollution Levels

With air quality improvements observed in other cities from restrictions on movement/industry that lead to significant emission reductions, changes in ambient background levels near Mukuru Kwa Reuben during lockdown were contextualized with probable significant increases in localized HAP levels from transition to polluting cooking fuels. A crude analysis of ambient air pollution fluctuations revealed the monthly mean ambient PM_2.5_ level (averaged across all publicly accessible laser scattering monitors surrounding the study setting) being nearly 50% lower in April 2020 (7 µg/m^3^) compared with the prior year (April 2019: 13 µg/m^3^). A comparison of daily average 24-hour PM_2.5_ levels revealed 12 days in April 2019 with average 24-hour PM_2.5_ concentrations greater than the WHO 24-hour ambient air quality guideline (25 µg/m^3^), while all daytime average PM_2.5_ concentrations in April 2020 were less than 13 µg/m^3^ (Figure 3). A clear downward trend in average 24-hour ambient PM_2.5_ levels around Mukuru Kwa Rueben after the lockdown in Kenya took effect (early April 2020) was observed (Figure S1).

**Figure 3:**
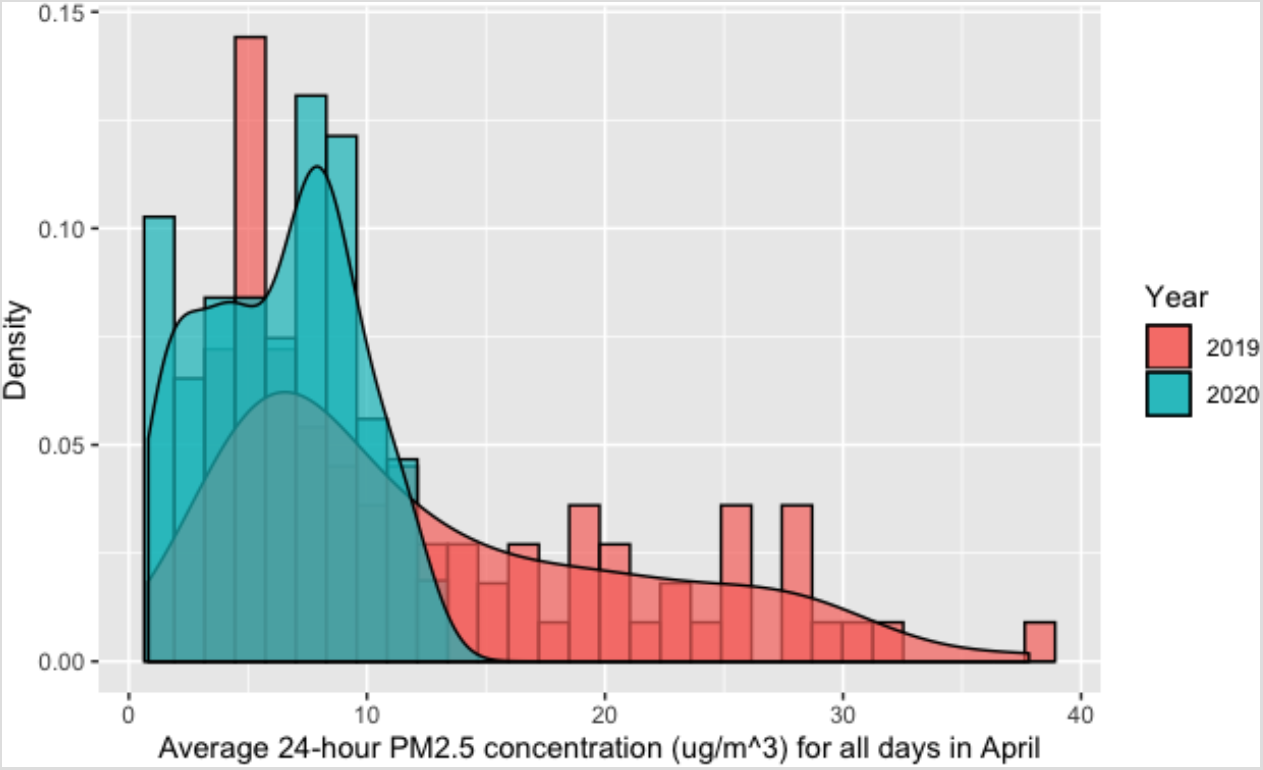
Comparison of 24-hour average ambient PM2.5 concentrations surrounding Mukuru Kwa Reuben informal settlement in April 2019 and April 2020

## DISCUSSION

This timely pre-post study documented an extremely high proportion (88%; n=180) of households experiencing food insecurity due to income decline/cessation during the COVID-19 lockdown in a Kenyan urban informal settlement (Table 3). As a result, 8% of families resorted to farming/livestock as their main/only source of food, and half (52%; n=101) switched their diets to commonly include higher intake of vegetables, as they were no longer able to afford meat/fish, milk and bread (Table S5), increasing the likelihood of protein deficiency.^37^ A decrease or loss of income during lockdown resulted in 27% of households switching their primary cooking fuel from clean LPG to a polluting fuel (kerosene, wood) (Table 3), as these fuels could be purchased daily for small sums of money (kerosene/wood) or gathered for free (wood). While LPG was reported as the least expensive fuel on a monthly basis (Table 1), the need to purchase a cylinder refill, and documented increase in the price of LPG compared with kerosene during lockdown (Table S8), likely played critical roles in families opting to purchase kerosene instead.

With 8% of households that primarily cooked with kerosene before the lockdown also switching to firewood while in lockdown, a 9% rise in prevalence of firewood cooking fuel among the study population (2% to 11%) (Figure 2) likely contributed to a rise in community-level HAP concentrations.^38^ Increased HAP exposure can be experienced by both the households where the switching to polluting cooking fuels occurred and nearby families, due to infiltration of biomass smoke to the surrounding area,^32^ especially with tightly-packed housing in an informal settlement.

Although 76% of families cooked less frequently during home lockdown, the length of HAP exposures may still increase, as meals prepared with fuelwood required an average daily cooking time that is 1.5 hours longer than with LPG (Table 1), and because families were confined inside the home in close proximity to the cooking area (89% of households had less than three rooms) (Table 1). Elevated HAP exposures among the study population may offset potential health benefits from the decrease in ambient air pollution levels during COVID-19 lockdown (Figure 3),^39^ which were documented in studies conducted in high-income countries.^40,41^ Increased PM_2.5_ exposure due to HAP is a risk factor for several chronic respiratory/cardiovascular health conditions, and may also increase the collective population’s vulnerability to suffering from more severe symptoms of COVID-19; recent studies have linked COVID-19 severity to increased chronic exposure to ambient air pollution.^42^

The limited number of households solely relying on electricity for lighting demonstrates its unreliability in the study setting; renewable energy cooking solutions (e.g. solar/grid electricity) are far from having the scalability needed to reach large sectors of the population.^43^ With over 90% of non-LPG users that had previous experience cooking with LPG expressing interest in future use (Table S10), LPG represents an encouraging clean energy source in informal settlements in Nairobi. However, to overcome the affordability barrier to LPG uptake expressed by 67% of baseline non-LPG users (Table S10), which may increase in the aftermath of the COVID-19 pandemic, there is an urgent need for consumer-centric approaches, including consumer-finance mechanisms (e.g. pay-as-you-go (PAYG), mobile payments) to offer families the opportunity to pay on an as-needed basis.^19^ The high cost of LPG refills (85% of LPG households used 6 kg cylinders) explains the appeal of small incremental payments via PAYG smart-meter technology among participants (Table 2). With the launch of two commercial companies in Nairobi that offer PAYG LPG, this innovation represents a promising solution for increasing LPG penetration in the study setting. Two-thirds of current kerosene users reported an intention to register with a PAYG LPG company (30%) or being interested in learning more about the technology (35%) (Table 2). A recent intervention study assessing the acceptability of PAYG payments for pellets (with gasifier stoves) in peri-urban Kenya found that PAYG displaced traditional fuels in many households, and participants had higher willingness to pay for PAYG technology, compared with non-PAYG stoves.^44^

Participants not interested in registering for PAYG LPG in the study community had higher average monthly incomes than the study population average, suggesting that the barrier of LPG affordability addressed by PAYG LPG was of less relevance to these households. Nevertheless, 52% of LPG primary users were interested in registering for PAYG LPG technology because of other advantages, including perceived increased safety (27%), customer service (13%) and the ability to monitor their LPG usage (12%).

Expanding the LPG market in Kenya and across SSA presents a viable, medium-term pathway to achieving universal clean household energy access (United Nation’s Sustainable Development Goal (SDG) 7).^19^ However, there are new challenges brought on by the COVID-19 pandemic: in addition to a lower purchasing power among low-income populations, a recent Clean Cooking Alliance survey of companies involved in fuel supply chains (stove/fuel manufacturers, distributors), NGOs and investors showed that 70% of enterprises had moderate/severe disruptions in their work activities, with 20% ceasing operations.^45^ Nearly two-thirds of clean cooking enterprises echoed significant concerns about reduced customer ability to pay for clean cooking fuels during the pandemic.

While this pre-post study had a lower-than-expected follow up rate (41%), primarily as a result of participants not willing to be re-contacted or discontinuing mobile phone plans due to financial hardship, a follow up sample of 194 participants, largely representative of the baseline population (Table S2), was powered to detect significant changes in food access and fuel use patterns during lockdown. The publicly available data from laser scattering PM_2.5_ monitors used in this study, while not gravimetrically corrected to improve accuracy, provide crude evidence of ambient air pollution fluctuations, particularly with concentrations aggregated across multiple sensors to minimize measurement error. While the socioeconomic and fuel pricing data is self-reported, the dramatic results provide strong evidence of the negative side-effects of confinement measures on the livelihoods of Africans largely working in the informal economy.^23^

While the COVID-19 pandemic presents a precarious situation for countries in SSA with weak healthcare systems,^46,47^ lockdown measures aiming to mitigate the spread of COVID-19 can intensify existing challenges to combat global hunger and gain momentum in the clean energy sector. There is a need for COVID-related interventions to be specifically tailored to the socioeconomic context to minimize unnecessary financial fallout.^34^ While the long term effects of the pandemic on consumers’ decisions regarding household energy are unclear, the cobenefits to health, climate and livelihoods should make universal access to clean household energy a priority for governments and global health leaders moving forward.

## Data Availability

Survey data collected as part of this study will be made available on a case-by-case basis on request to the corresponding author, with input from the co-authors, subject to compliance with Research Ethics Board restrictions.

## Acknowledgements

The authors would like to sincerely thank the field team in Mukuru Kwa Reuben for their flexibility in transitioning from in person to telephone-based surveys. Without their extensive efforts and dedication to research during the COVID-19 pandemic, this study would not be possible.

## Author contributions

MS, EP, DP and JM designed the study. AG and JM led the field work. MS wrote the paper and carried out the analysis. All co-authors discussed the results and reviewed the manuscript.

